# Identification of Health Conditions in Unstructured Health Records with Deep Learning-Based Natural Language Processing

**DOI:** 10.1101/2024.10.08.24315141

**Authors:** Jake Lin, Tomi Korpi, Anna Kuukka, Anna Tirkkonen, Antti Kariluoto, Juho Kaijansinkko, Maija Satamo, Hanna Pajulammi, Markus J. Haapanen, Sergei Häyrynen, Eetu Pursiainen, Daniel Ciovica, Mikaela B. von Bonsdorff, Juulia Jylhävä

## Abstract

**Importance:** Many clinically significant health conditions are frequently underreported, underdiagnosed or recorded only in unstructured textual health records, yet they contain critical information for patient assessment, care and prognosis.

**Objective:** To determine whether deep learning-based natural language processing employed for named entity recognition can effectively identify health conditions, such as incontinence, falls, mobility limitations and loneliness in unstructured textual electronic health records. The identified conditions were further used to predict all-cause mortality.

**Design:** This cohort study utilized electronic health records from public primary, secondary, tertiary, long-term and home care from 2010 to 2022, providing up to 12 years of follow-up. The named entity recognition task to identify incontinence, falls, mobility limitations, and loneliness was implemented using Google’s Bidirectional Encoder Representations from Transformers deep learning model pre-trained for the Finnish language. Diagnostic codes for incontinence and falls were collected for comparisons.

**Setting:** Retrospective electronic health records across the Central Finland wellbeing services county.

**Participants:** Structured summary data and 10.6 million free-text entries from 102,525 patients aged 50 to 80 years at baseline.

**Exposure:** Incontinence, falls, mobility limitations and loneliness were considered as exposures.

**Main Outcomes and Measures:** The performance of the named entity recognition models was evaluated by precision, recall and F1 scores benchmarked against human ratings. Cox regression models were used to assess and compare NER- and diagnostic code-identified falls and incontinence onsets in predicting all-cause mortality.

**Results:** The deep learning model demonstrated excellent performance with recall, precision and F1 scores of 0.86, 0.88, and 0.87 for falls; 0.84, 0.78, and 0.81 for incontinence; 0.86, 0.84, and 0.85 for mobility limitations and 0.91, 0.84, and 0.87 for loneliness, respectively. Compared to diagnostic codes, named entity recognition identified greater numbers of falls (31987 vs 4090) and incontinence (7059 vs 3873) onsets and yielded greater hazard ratios: 1.31 vs 1.04 for falls and 1.99 vs 0.65 for incontinence.

**Conclusions and Relevance:** Deep learning-based named entity recognition models reliably identified incontinence, falls, loneliness and mobility limitations in free-text medical records, presenting new opportunities to use unstructured clinical data to identify vulnerable patients and apply the method in research applications.

**KEY POINTS:** *Question:* Can deep learning-based natural language processing (NLP) identify health conditions, such as incontinence, falls, mobility limitations and loneliness in unstructured electronic health records (EHRs)?

*Findings:* The results of this cohort study demonstrate that a deep-learning NLP model can effectively identify incontinence, falls, mobility limitations and loneliness in textual EHR data. This approach also results in improved mortality prediction compared to available diagnostic codes.

*Meaning:* NLP approaches could be used to identify underreported and underdiagnosed health conditions in textural EHR data, enabling identification of vulnerable and at-risk patients.

## INTRODUCTION

Many clinically significant conditions in older patients, such as incontinence, falls, mobility limitations and loneliness are frequently underreported, underdiagnosed or recorded only in unstructured data within electronic health records (EHRs).^1–3^ Data on these conditions nevertheless provide relevant information on individual’s health and functioning beyond diagnostic disease codes and hold significant value for more detailed patient assessment, care planning and prognostic purposes. Until recently, identifying and extracting such information from the free-text EHRs has been inaccessible on a larger scale as traditional methods, such as manual abstraction are insufficient for processing unstructured text efficiently. Modern techniques, such as artificial intelligence (AI) -guided natural language processing (NLP) have been developed to process and interpret large quantities of unstructured textual data,^4,5^ capturing previously inaccessible information on individual’s health and functioning.

NLP is a broad field encompassing a variety of techniques designed to understand, interpret, and generate human language.^4,5^ Despite advancements in NLP, there is limited knowledge whether specific NLP applications, such as named entity recognition^6^ (NER) can accurately identify health conditions from unstructured EHR data. Challenges include designing effective clinical NER systems with large quantities of representative training data to ensure generalizability and domain-specific performance. Data on the precision and reliability of these models across different clinical settings is also limited.^7^ NER applications that use deep learning techniques have however demonstrated potential in extracting relevant clinical entities from large volumes of text.^7^

This study was undertaken to analyze whether a deep learning-based NLP model using NER and Google’s Bidirectional Encoder Representations from Transformers (BERT)^8^ can effectively identify commonly underreported and underdiagnosed age-related health conditions, namely incontinence, falls, mobility limitations and loneliness in unstructured EHR data. We also compare the onsets of falls and incontinence identified by the NER model to those derived using diagnostic codes (International Classification of Diseases, 10^th^ Revision; ICD-10) and assess the performance of the NER vs ICD-10-based models in predicting all-cause mortality. We hypothesize that our NER approach to free-text EHRs would identify a greater number of at-risk individuals compared to ICD-10 codes and lead to improved risk stratification of mortality.

## METHODS

### Sample

All inpatient and outpatient EHRs from public primary, secondary, tertiary, long-term and home care across the Central Finland wellbeing services county using the Lifecare EHR system between 2010 and 2022 were used in the analysis. The records included ICD-10 codes, free-text entries and routine laboratory tests. The free-text entries, recorded by healthcare professionals, included all recorded clinical notes (e.g., progress, admission/discharge summaries, operative reports, consultation notes), physical examination notes, nursing notes, physiotherapy and rehabilitation notes, psychiatric notes, radiology reports, care plan notes and patient and family communication, including telephone encounters. The dataset included 10.6 million free-text entries from 102,525 patients, with baseline ages (i.e., age at first EHR entry) ranging from 50 to 80. For ethical considerations, see eMethods (Supplement 1).

### Named entity recognition

The EHR data were de-identified of personal information and extracted with relevant keywords/phrases for each condition i.e., falls, incontinence (including both urinary and fecal incontinence), loneliness and mobility limitations. Subsequently, subsampling (10-15% of the full sample) was performed to create datasets for manual labelling. FinBERT^9^, an extension of a deep neural network-based BERT model to the Finnish language with a custom NER approach was implemented to identify the onsets (i.e., first occurrences) of falls, incontinence, loneliness and mobility limitations in the free-text EHR data. For falls, incontinence and loneliness, we considered the conditions as binary (yes/no), whereas for mobility limitations, we labeled the text entries in two categories: 1) mobility limitations e.g. person being able to ambulate independently but with some limitations or pain, and 2) more severe mobility limitations with the person needing personal assistance or wheelchair and/or mobility aids with ambulation or is non-ambulatory. A detailed description of our NER pipeline and examples of passages used to identify the health conditions are provided in eMethods, eTable 1 and eFigure 1 (Supplement 1).

**Figure 1.**
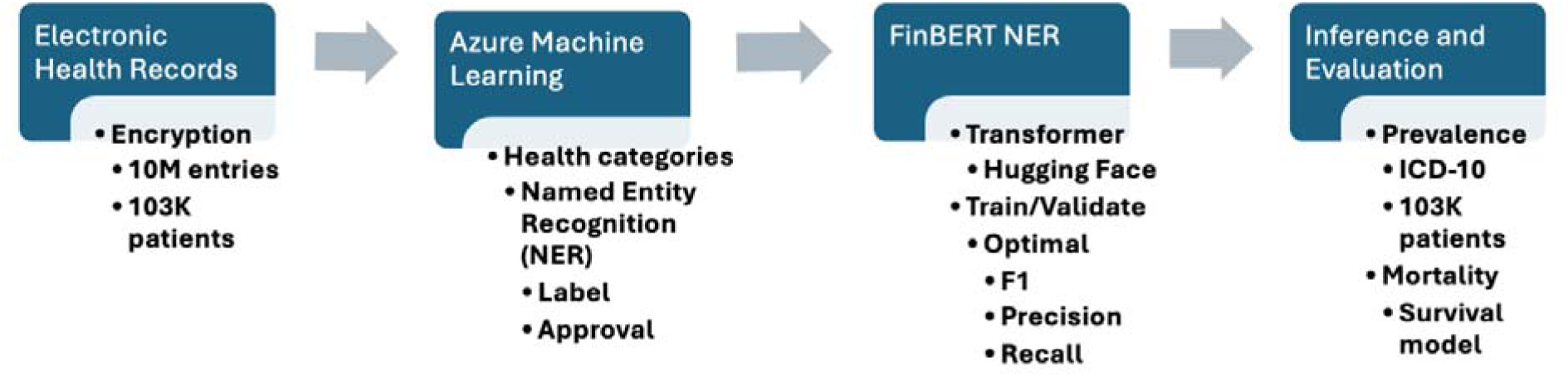
Workflow of the study. Textual electronic health records (EHRs) were encrypted and scrubbed of all personal information. Working with domain experts, we applied keywords to identify relevant EHR entries and performed appropriate subsampling for labelling within Azure Machine Learning Studio. FinBERT library, a pretrained BERT model adopted for the Finnish language, and the Hugging Face-associated transformer libraries were integrated for the processing, training, and evaluation of the labelled and approved EHR text data. Abbreviations: ICD-10, International Classification of Diseases, 10^th^ Revision; NER, named entity recognition

Relevant texts were labelled for each condition by a domain expert and subsequently corrected as necessary and approved by another domain expert. Assisted by clinicians and nurses, the labelers and approvers were mutually exclusive relative to all EHR NER entries. Absence of a deficit in the patient’s text entries was considered as no deficit i.e., only positive text entries were labeled. However, pertinent to loneliness, to address the known negation limitations in BERT^10^, the identification of negative terms (no experiences of loneliness) was additionally performed with a custom application of Levenshtein distance^11^ as implemented in Python.

Upon transformation to the BIO labelling format required for the NER model, the dataset was split into 80/10/10 proportions for training, validation and testing. Tokenization mapping was performed on the original text where numeric values were aligned to the relevant BIO labels. Taking advantage of the open-sourced Hugging Face transformer^12^ and evaluation libraries, training was performed. For each health condition and corresponding model, the pivotal hyper parameters, learning rate and weight decay were tuned as part of a custom and iterated evaluation function. The optimal models, based on F1 value (equation 3 below), were saved and subsequently applied for the inference step. The inference transformer pipeline performs token classification and evaluation on all EHR entries of the listed health categories, calculating confidence scores on each custom entity and relevant labels. The workflow of the study is presented in Figure 1.

### Infrastructure

NER data labelling and approval were performed on the Microsoft Azure Machine Learning studio program, provided by the secured and GPU-enabled cloud infrastructure of the Helsinki and Uusimaa Hospital District, required for the underlying CUDA cores necessary for parallel computing driving deep learning in transformer models.

### Performance

The overall statistical performance of NLP learning classifiers was appraised, as defined with the accompanied equations, with the standard aid of the precision (1), recall (2) and their harmonic mean F1 score (3) parameters. Strict and partial matches were compared with their performance metrics defined by true positive (TP) and true negative (TN), false positive (FP) and false negative (FN) designations with respect to TP, as assigned, and relative to expert manual human predictions within the produced confusion matrix. We targeted an overall threshold F1 score of 0.80, generally accepted to be excellent. The equations of the evaluation parameters are:

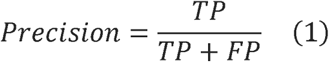

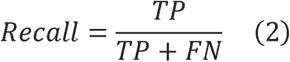

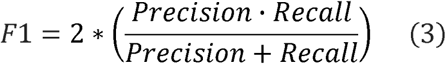

### Inference

Inference was performed with the persisted model towards matched records extracted from the associated original keywords. An inference token probability score,^13^ provided by the Hugging Face inference pipeline^12,14^ of at least 0.90 is applied to discriminate similarity and avoid false positives. Additionally, the numbers of onsets of the conditions from the NER models were compared to ICD-10-based onsets for falls and incontinence at the individual level. ICD-10-based falls were defined by codes W00-W19 (excluding jumping into water/pools W16.5-W16.9), while incontinence was defined by codes N39.4, R15, R32 and R39.81. Processing of the scripts was accomplished with Jupyter Python (3.09) notebooks, while inferences modeling was conducted in R version 4.4.0.

### Statistical analyses

We performed Cox regression models on all-cause mortality to contrast differences in the predictive strengths of NER- and ICD-10-based falls and incontinence (ICD-10 codes do not exist for mobility limitations and loneliness). The onset of each condition was defined at the first occurrence of the condition (model baseline). Individuals without an event (death) were censored at the end of the follow-up 31^st^ December 2022. The models, adjusted for attained age and sex, were performed in the full sample and stratified by sex. Statistical analyses were performed using R software version 4.4.0. Hazard ratio (HR) and its 95% confidence interval (CI) were computed using ‘coxph’ from the survival package. P-values <0.05 were considered statistically significant.

## RESULTS

Our dataset included 102,525 (51.4% female) patients with 10.6 million valid free-text entries collected between the 1^st^ of January 2010 and 31^st^ of December 2022. During the collection period, 21,213 (20.7%) patients had died, with a mean age of 76.2 and standard deviation (SD) 8.7 at baseline. Participant details are shown in Table 1.

**Table 1.** Characteristics of participants in the electronic health record data.

| Characteristics |  |
| --- | --- |
| N Individuals | 102525 |
| N Free-text entries, total | 10.6 million |
| N Free-text entries, per person mean (SD) | 103.8 (199.5) |
| Age at baseline, mean (SD) | 63.84 (8.09) |
| Age at baseline, min-max | 51.00-80.92 |
| Sex |  |
| N Male (%) | 49883 (48.65) |
| Age at baseline, mean (SD) | 63.57 (7.94) |
| Age at baseline, min-max | 51.00-80.92 |
| N Female (%) | 52642 (51.35) |
| Age at baseline, mean (SD) | 64.11 (8.21) |
| Age at baseline, min-max | 51.00-80.92 |
| N Died between 2010 and 2022 (%) | 21213 (20.69) |
| Age at death, mean (SD) | 76.20 (8.68) |
| Age at death, min-max | 51.10-93.00 |
| Time (years) to follow-up*, median (IQR) | 11.41 (11.08-11.75) |
| Time (years) to death, median (IQR) | 7.00 (3.58-9.83) |

\*Time to follow-up includes all participants, also those who died.
Abbreviations: IQR, interquartile range; SD, standard deviation

The Jupyter Python pipeline, featuring configuration-based processing, hyperparameter default values and finetuning results are shown in eMethods, eTable2 and eTable 3, respectively (Supplement 1). The deep learning FinBERT-based NER models demonstrated excellent performance with recall, precision, and F1 scores of 0.86, 0.88, and 0.87 for falls; 0.84, 0.78, and 0.81 for incontinence; 0.91, 0.84, and 0.87 for loneliness and 0.86, 0.84, and 0.85 for mobility limitations (independent with limitations and requiring personal assistance -categories pooled). The corresponding results using a strict evaluation are shown in eTable 4 (Supplement 1). As shown in Table 2, the NER model identified significantly more falls (31987 vs 4090) and incontinence (7059 vs 3873) onsets. There was a significant overlap between the ICD- and NER-identified onsets, such that the NER model identified 90.1% of fall and 31.0% of incontinence onsets identified by ICD-10 codes (Table 2). Based on our NER models, we found that the proportions of patients with falls, incontinence, loneliness and mobility limitations increased with respect to age, with the highest onset proportions in the oldest (aged 80+) populations (Figure 3). The numbers and proportions of individuals having the conditions by age category are presented as numeric in eTable 5 (Supplement 1). The overlap of the conditions among the patients identified by the NER model is presented in eFigure 2 (Supplement 1), showing a degree of overlap across the conditions but also a significant number of patients with only one condition.

**Figure 2.**
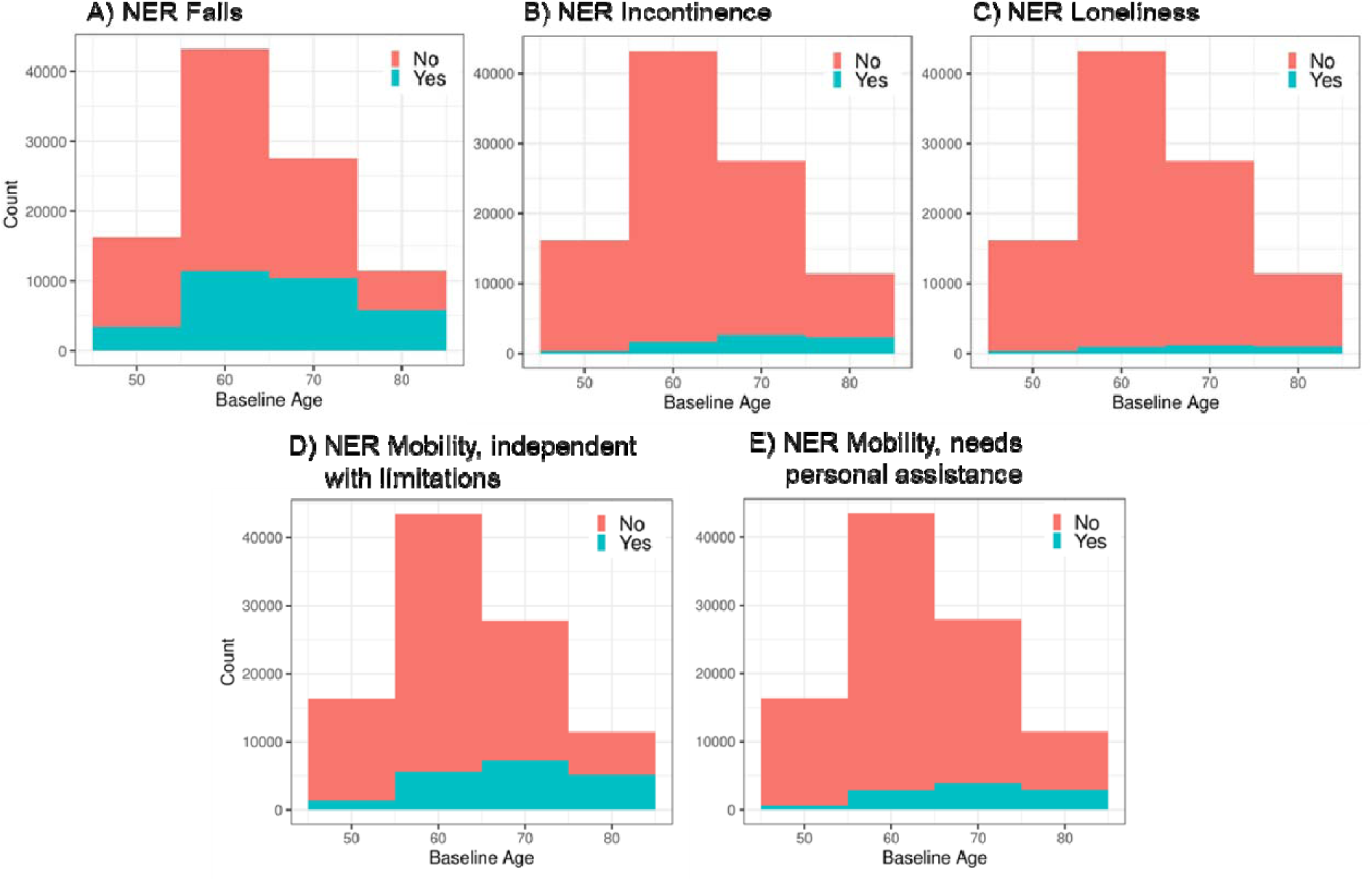
Numbers of falls (A), incontinence (B), loneliness (C) and mobility limitations (D and E) based on the named entity recognition (NER) models by age group. Baseline age defines the first occurrence of the conditions.

**Table 2.** Precision, recall and F1 scores for falls, incontinence and mobility limitations based on optimal settings and partial approximation evaluation matching. The intersection percentage is relative to the number of ICD-10 identified onsets that are in common with the NER model.

| Health condition | Labeling N | F1 | Recall | Precision | Onsets by NER (Female %) | Onsets by ICD-10 (Female %) | NER ICD-10 Intersection (%) |
| --- | --- | --- | --- | --- | --- | --- | --- |
| Falls | 3000 | 0.87 | 0.86 | 0.88 | 31987 (56.4) | 4090 (59.3) | 3639 (90.1) |
| Incontinence | 3000 | 0.81 | 0.84 | 0.78 | 7059 (57.4) | 3873 (79.4) | 1200 (30.0) |
| Loneliness | 1000 | 0.87 | 0.91 | 0.84 | 3623 (67.7) | NA | NA |
| Mobility limitations | 5000 | 0.85 | 0.86 | 0.84 | 21690 (55.1) | NA | NA |

Note. The labelling N indicates the number of free-text entries used in training the model. Abbreviations: ICD-10, International Classification of Diseases, 10th Revision; NER, named entity recognition

As shown in Table 3, the NER-based Cox models of falls and incontinence on all-cause mortality were more predictive compared to the ICD-10-based models. After adjusting for attained age and sex, the HRs and their 95% CI for NER-based falls in the full sample were 1.31 (95% CI 1.27-1.35), compared to ICD-10-based HR of 1.04 (95% CI 0.99-1.10) (Table 3). With respect to the NER-based survival model of incontinence in the full sample, we found a HR of 1.99 (95% CI 1.92-2.06), compared to a lower HR of 0.65 (95% CI 0.61-0.70) for the ICD-10-based model (Table 3). We investigated the unexpected HR for ICD-10-based incontinence and found that 78% of individuals with incontinence onset defined by ICD-10 were still alive, explaining the HR in the opposite direction of expectations. The NER-based survival models of mobility, independent with limitations (HR 1.89; 95% CI 1.84-1.96) and mobility limitations, needing personal assistance (HR 2.19; 95% CI 2.12-2.26) were predictive of mortality. However, results for the NER-based survival model of loneliness (HR 0.97; 95% CI 0.92-1.03) were not predictive. The results across the models were consistent when stratified by sex (Table 3).

**Table 3.** Cox regression models on all-cause mortality using named entity recognition-based onsets of falls, incontinence, loneliness and mobility limitations and International Classification of Diseases, 10th Revision-based onsets of falls and incontinence.

| NER-based model |  |  |  | ICD-10-based model |  |  |  |
| --- | --- | --- | --- | --- | --- | --- | --- |
| Health condition | HR | 95% CI | p |  | HR | 95% CI | p |
| **Falls** |  |  |  |  |  |  |  |
| Full sample | 1.31 | 1.27-1.35 | p<2e-16 |  | 1.04 | 0.99-1.10 | 0.14 |
| Men | 1.39 | 1.34-1.44 | p<2e-16 |  | 1.12 | 1.03-1.21 | 0.006 |
| Women | 1.20 | 1.15-1.25 | p<2e-16 |  | 0.98 | 0.90-1.05 | 0.52 |
| **Incontinence** |  |  |  |  |  |  |  |
| Full sample | 1.99 | 1.92-2.06 | p<2e-16 |  | 0.65 | 0.61-0.70 | p<2e-16 |
| Men | 2.02 | 1.92-2.12 | p<2e-16 |  | 0.61 | 0.56-0.67 | p<2e-16 |
| Women | 1.95 | 1.85-2.05 | p<2e-16 |  | 0.74 | 0.66-0.92 | 0.09 |
| **Loneliness** |  |  |  |  |  |  |  |
| Full sample | 0.97 | 0.92-1.03 | 0.37 |  | - | - | - |
| Men | 0.95 | 0.87-1.04 | 0.30 |  | - | - | - |
| Women | 0.98 | 0.91-1.06 | 0.66 |  | - | - | - |
| **Mobility, independent with limitations** |  |  |  |  |  |  |  |
| Full sample | 1.89 | 1.84-1.96 | p<2e-16 |  | - | - | - |
| Men | 1.96 | 1.89-2.05 | p<2e-16 |  | - | - | - |
| Women | 1.80 | 1.72-1.88 | p<2e-16 |  | - | - | - |
| **Mobility, needs personal assistance** |  |  |  |  |  |  |  |
| Full sample | 2.19 | 2.12-2.26 | p<2e-16 |  | - | - | - |
| Men | 2.24 | 2.15-2.34 | p<2e-16 |  | - | - | - |
| Women | 2.12 | 2.02-2.22 | p<2e-16 |  | - | - | - |

Note. The models were adjusted for attained age and sex when applicable. ICD-10-based falls were defined by codes W00-W19 (excluding jumping into water/pools W16.5-W16.9), while ICD-10-based incontinence were defined codes N39.4, R15, R32 and R39.81. Abbreviations: CI, confidence interval; HR, hazard ratio, ICD-10, International Classification of Diseases, 10<sup>th</sup> Revision; NER, named entity recognition

## DISCUSSION

Approximately 80% of medical records are unstructured textual data.^15^ These data require minimal maintenance yet contain a wealth of detailed information not captured in structured data, such as diagnostic codes. Diagnostic codes are also known to be underused, with systematic underreporting particularly prevalent in syndromes of older patients compared to other conditions.^16^ We present a deep learning AI-enhanced NLP approach using NER in Finnish language to extract such information on clinically relevant aging-related health conditions, namely falls, incontinence (both urinary and fecal), loneliness and mobility limitations in a large dataset of 10.6 million textual EHRs from primary, secondary, tertiary and institutional care, covering a period of 12 years.

Benchmarked against human ratings, our NER model demonstrated excellent performance with recall, precision, and F1 scores of 0.86, 0.88, and 0.87 for falls; 0.84, 0.78, and 0.81 for incontinence; 0.91, 0.84, and 0.87 for loneliness, and 0.86, 0.84, and 0.85 for mobility limitations. Compared to diagnostic codes, the NER models identified greater numbers of falls (31987 vs 4090) and incontinence (7059 vs 3873) onsets. The numbers of the NER-identified onsets of falls, incontinence, loneliness and mobility limitations increased with age as expected, demonstrating that our NER model is consistently effective across various age-related conditions. In predicting all-cause mortality, we found higher HRs for models of falls and incontinence when identified using NER compared to ICD-10 codes of falls and incontinence, supporting the fact that the ICD codes for falls and incontinence are underreported in structured summary data. Mobility limitations identified using NER were also predictive of mortality with higher HRs observed for more severe limitations (mobility needing personal assistance compared to independent with limitations), demonstrating that the NER model effectively captures a dose-response relationship between mobility limitations and mortality risk. Mortality prediction results for the loneliness NER model were not significant.

Different NLP systems relying on rule-based, machine learning and deep learning models have been previously used to identify and classify age-related conditions, such as sarcopenia, limitations in activities of daily living and falls.^17,18^ These models have yielded F1 scores ranging from 0.53 to 0.95, though several of the studies have not reported any evaluation metrics.^17,18^ The performance of our NER models (all F1 scores >0.80) thus falls at the high end of the F1 scale, although exact comparison is difficult due to differences in the approaches and data-related characteristics, such as the size of the training data and types of notes and patient groups included. It is noteworthy that most studies using NER have used public data sets, while only a minority (7%) of the models have had access to private data requiring exclusive licenses or external permissions.^7^ Use of large private, real-world EHR data from diverse patient groups, such as in our study, enhances generalizability and reduces bias, resulting in models that are adaptable to various clinical settings.^7^ Most previous NER studies have also used English or Chinese corpora,^7^ highlighting the need for more research in other languages. Our study, utilizing a Finnish language model, addresses this gap and demonstrates the ability of a FinBERT^9^ NER model to perform clinical NLP tasks.

Although our NER model identified more fall and incontinence onsets compared to their corresponding ICD-10 codes, supporting the known fact that structured data -based reports tend to underreport health outcomes,^16^ the overlaps between the NER and ICD-10-identified onsets were significant but only partial: 90.1% for falls and 31.0% for incontinence. The incomplete overlap may result from the absence of any mention of a fall or incontinence in the text, or from misclassification by the model. Given the high recall values i.e., true positive rates for the NER-identified falls and incontinence, misclassification is unlikely to explain the discrepancy. Affirming the validity of our NER models, the numbers of the NER-identified falls, incontinence, loneliness, and mobility limitations followed an expected age-associated pattern, with the highest proportions observed in the oldest populations (aged 80+). This trend aligns with existing literature, which demonstrates that the prevalence of these conditions increases with advancing age.^19,20^ Comparison the numbers of the onsets to previous studies using NLP in similar multi-specialty settings as ours reveals comparable estimate ranges for falls, incontinence and mobility limitations.^17,18,21^ Several clinical NLP studies have however used only specific types of clinical notes or focused on a distinct medical specialty or patient group,^7,17^ making comparisons infeasible. For example, the proportion of patients suffering from loneliness has previously been assessed only in patients receiving mental healthcare,^22^ resulting in higher prevalence estimates (16.7%) compared to our setting (2.0-11.2%) that included records across all medical specialties.

Our NER-based models of falls and incontinence were more predictive of all-cause mortality compared to ICD-10-based models, indicating that NER-based models provide a more accurate assessment of mortality risk, which is crucial for effective clinical management and intervention. The ICD-10 codes for falls are typically recorded in the context of injurious falls, which may not be predictive of long-term mortality. In contrast, ostensibly uninjurious falls, such as those occurring in nursing homes, are only recorded in free-text records; these falls may also indicate recurrent falls and thus better capture the risk of mortality. Capturing such falls is nevertheless of utmost importance, for example for fall prevention, as previous falls have been identified as the strongest predictor of future falls.^23^ Loneliness based on the NER model did not however predict mortality, which is likely attributable to the fact that our model could not discriminate between transient and chronic loneliness. A recent study has shown that cumulative loneliness is predictive of mortality, whereas transient loneliness is less strongly or not at all associated with mortality.^24^ Notably, mobility limitations identified by NER likewise predicted mortality with higher HRs for greater degrees of limitation, yet the performance of these models could not be compared to ICD-10-based models as no corresponding diagnostic codes exist. The estimates were highly similar for men and women across all NER models, suggesting no sex bias in the utility of the free-text information. Overall, the results demonstrate that free-text information on these health conditions can be used to better identify high-risk patients and support decision-making regarding care.

### Limitations

The limitations of our study are pertinent to all studies using EHR data; the data can be incomplete, with gaps in clinical documentation, information can be missing or the entries are incorrect, affecting the accuracy of the models. EHRs are also often filled in by different healthcare providers, leading to inconsistencies in how information is recorded. Misspellings and abbreviations may also pose difficulties for NLP models to interpret the data. Although our data were sourced from one wellbeing services county in Finland, the sample is expected to represent the general Finnish population, yet we did not have an external validation sample available to test the generalizability of the results. The large size of our sample (10.6 million text records) nevertheless ensured a sufficient amount of data for training the models – an aspect known to increase generalizability.

## CONCLUSIONS

With 10.6 million EHR texts, our study is among the largest to date to demonstrate the utility of clinical NLP models in identifying health conditions in real-world patient data. With diverse multi-specialty training data, our analysis was not limited to distinct patient groups or healthcare units, which is expected to enhance generalizability. Importantly, our results support the use of the BERT architecture for clinical NER tasks in the Finnish language to identify falls, incontinence, loneliness, and mobility limitations. Overall, our results add to the existing literature on NLP-identified health issues carrying clinically relevant information that can be used to identify vulnerable, high-risk patients and support healthcare decision-making.

## Data Availability

Electronic health records used in this data are personally identifying and hence not publicly available. Our code and pipelines implemented as Python Jupyter notebook scripts are available at: https://github.com/jakelin212/frailty_nlp_ner

## Author contributions

Dr Lin had full access to all of the data in the study and takes responsibility for the integrity of the data and the accuracy of the data analysis.

**Concept and design:** Jylhävä, Lin, Häyrynen.

**Acquisition, analysis, or interpretation of data:** Lin, Korpi, Kuukka, Tirkkonen, Kariluoto, Kaijansinkko, Satamo, Haapanen, Pajulammi, Häyrynen, Pursiainen, Ciovica, von Bonsdorff, Jylhävä.

**Drafting of the manuscript:** Lin, Jylhävä, Korpi.

**Critical revision of the manuscript for important intellectual content:** Lin, Korpi, Kuukka, Tirkkonen, Kariluoto, Kaijansinkko, Satamo, Haapanen, Pajulammi, Häyrynen, Pursiainen, Ciovica, von Bonsdorff, Jylhävä.

**Statistical analysis:** Lin, Kuukka, Kariluoto.

**Administrative, technical, or material support:** - **Supervision:** Lin, Jylhävä.

## Data sharing statement]

See Supplement 2.

