## Supplemental Figures and Table for "Identification of Health Conditions in Unstructured Health Records with Deep Learning-Based Natural Language Processing"

**Supplement**

**Supplementary methods**

*Ethical considerations*

Research permit to use the data was obtained from the central Finland wellbeing services county. Informed consent and review and/or approval by an ethics committee were not required for this study as according to the Finnish legislation (Act on the Secondary Use of Health and Social Data (552/2019) by the Ministry of Social Affairs and Health), the patients included in the study sample were not contacted, and the study did not affect the treatment of the patients. The Human Sciences Ethics Committee of the University of Jyväskylä has certified these conditions pertinent to our study and stated that an ethical review is not required. The study was conducted in accordance with the Helsinki Declaration.

*Named Entity Recognition*

Named entity recognition (NER) is a common and necessary task to process and standardize the unstructured text using Natural Language Processing (NLP) techniques to extract relevant information from the data. It involves managing labeling projects to identify and assign entities to categories in the unstructured text by manual labeling, in our case, in the Azure ML Studio. The number of subsampled assets applied were in the range of 1500-5000 (approximately 10-15% of the relevant health record entries based on comprehensive list of keywords). Supplementary Table 1 below outlines example text sequences based on the most common expressions for each deficit, translated from Finnish, pertaining to each health deficit category as presented.

The NER workflow, using the FinBERT transformer model is demonstrated in Supplementary Figure 1, showing the architecture of the NER task using pre-trained transformer models. The NER processing task is formulated as a sequence labeling task, to assign a predefined B/I/O tag[1] to each word (token) of the string sequence, where "B" means the beginning of an entity, "I" represents tokens inside a sequence, and "O" represents all other nonentity (outside) words. At first, the annotated texts in each asset were preprocessed and transformed into the "BIO" format (e.g., sentence boundary detection and initial tokenization), then the input instances were processed by appending with a special token [CLS] at the beginning of the sequenced text. The processed inputs were tokenized based on the pre-trained FinBERT[2] (Finnish based BERT shown to outperform BERT for Finnish based text) model's vocabulary and then fed into the language model where the contextual representations of the tokenized processed input were processed. Finally, the NER task is done by using an additional classification layer on the contextual representations to predict token tags. All transformer models were downloaded from the HuggingFace website (https://huggingface.co/models)[3] and the models were trained using the Transformers library as implemented by the HuggingFace team utilizing PyTorch (version 3.10). Due to the large size and sensitivity of our data, the entire processing workflow (sampling, labeling, training, evaluation and inference) are performed on HUS secure server environment (https://hustietoallas.fi) where GPU resources are available. Our parameter-based pipelines implemented as Python Jupyter notebook scripts are hosted at: https://github.com/jakelin212/frailty_nlp_ner:

For each health category, a tenfold cross-validation (train/validation/ testing subsets with a ratio of 80%:10%:10%) was used to train and evaluate the performance of the NER models. Based on literature[4] and recommendations,[5] the training hyperparameters as shown in Supplementary Table 2, with the exception of iteratively tuned (based on F1 trajectories) learning rate and weight decay were used for all the models. We evaluated the performance of all the transformer-based NER models using both the strict and partial precision, recall, and

F1-score,[6] where strict means that an entity is correctly identified if both the boundary and entity type is exactly as those in the standard (human experts manually annotated/approved in Azure Labeling). The partial mode assumes that an entity is correctly identified, if its entity type is correct and if its boundary overlaps with that in the human approved assigned mappings. As indicated in Supplementary Table 3 with its optimal tuned hyperparameters learning rate and weight decay, we found that for our health conditions, the partial mode outperforms the strict mode (Supplementary Table 4). For each condition, the prevalence rate and their onset ages, in spans of < 60, 60-70, 70-80 and 80+ years old are shown in Supplementary Table 5.

**Supplementary tables**

**Supplementary Table 1.** Example electronic health record text sequences relevant to our named entity recognition workflow. Tagged text are marked in bold.

| **Health deficit** | **Text sequences** |
|---|---|
| Falls | ‘patient had a **fall**’, ‘patient **fell**’, ‘patient found **fallen** on the floor, ‘patient **tripped** on to something’, ‘patient has had multiple **falls**’ |
| Incontinence | ‘patient is experiencing **loss of bladder/bowel control**’, ‘patient is suffering from **urinary/fecal incontinence**’, ‘patient experiencing **urinary/fecal leakage**’, ‘patient is **unable to withhold urine/stool**’, patient is **unable to control urination/defecation**’ |
| Loneliness | ‘patient reports feeling **lonely**’, ‘patient reports feelings of **loneliness**’, ‘patient suffers from **loneliness**’, ‘patient is experiencing **loneliness**’ |
| Mobility limitations, independent with limitations | ‘patient is **limping**’ ‘**moving triggers pain**’, ‘patient uses **walking aids [cane/walker/crutches/rollator]**, ‘patient needs to use **walls/handrails for support when walking**’, |
| Mobility limitations, needs personal assistance or e.g. wheelchair | ‘patient needs **personal assistance in ambulation**’, patient can only **move/walk with assistance by someone’**, ‘patient needs **a walker**, ‘patient uses/needs a **wheelchair**’, ‘patient needs **transfer aids**’, ‘patient is **bedbound/non-ambulatory/immobile in bed**’ |

**Supplementary Table 2.** Hyperparameters best practiced default values applied for all the transformer models.

| **Parameter** | |
|---|---|
| Batch size | 8 |
| Epoch | 10 |
| Learning rate | 1e-05 |
| Load best model | True |
| Maximum sequence length | 512 |
| Save steps | 100 |
| Weight decay | 0.2 |

**Supplementary Table 3.** Learning rate and weight decay hyperparameters for each health model were iteratively, as indicated by k, tuned for optimal F1 scores (indicated with an asterisk).

| Model | Learning rate | Weight decay | F1 score | Precision | Recall | k | Target |
|---|---|---|---|---|---|---|---|
| **Falls** | 1.0E-05 | 0.2 | 0.865* | 0.88 | 0.86 | 0 | Learning rate |
| | 2.0E-05 | 0.2 | 0.837 | 0.81 | 0.87 | 1 | Learning rate |
| | 3.0E-05 | 0.2 | 0.839 | 0.81 | 0.87 | 2 | Learning rate |
| | 4.0E-05 | 0.2 | 0.841 | 0.81 | 0.87 | 3 | Learning rate |
| | 5.0E-05 | 0.2 | 0.845 | 0.82 | 0.87 | 4 | Learning rate |
| | 6.0E-05 | 0.2 | 0.838 | 0.83 | 0.84 | 5 | Learning rate |
| | 7.0E-05 | 0.2 | 0.849 | 0.86 | 0.84 | 6 | Learning rate |
| | 8.0E-05 | 0.2 | 0.843 | 0.85 | 0.84 | 7 | Learning rate |
| | 1.0E-05 | 0.2 | 0.853 | 0.85 | 0.86 | 0 | Weight decay |
| | 1.0E-05 | 0.20001 | 0.857 | 0.85 | 0.86 | 1 | Weight decay |
| | 1.0E-05 | 0.20002 | 0.857 | 0.85 | 0.87 | 2 | Weight decay |
| | 1.0E-05 | 0.20003 | 0.86 | 0.86 | 0.86 | 3 | Weight decay |
| | 1.0E-05 | 0.20004 | 0.861 | 0.86 | 0.86 | 4 | Weight decay |
| | 1.0E-05 | 0.20005 | 0.856 | 0.84 | 0.87 | 5 | Weight decay |
| | 1.0E-05 | 0.20006 | 0.857 | 0.84 | 0.87 | 6 | Weight decay |
| | 1.0E-05 | 0.20007 | 0.864 | 0.86 | 0.86 | 7 | Weight decay |
| | 1.0E-05 | 0.20008 | 0.859 | 0.85 | 0.87 | 8 | Weight decay |
| | 1.0E-05 | 0.19999 | 0.86 | 0.85 | 0.87 | 9 | Weight decay |
| | 1.0E-05 | 0.19998 | 0.856 | 0.84 | 0.87 | 10 | Weight decay |
| **Incontinence** | 9.0E-05 | 0.19998 | 0.696 | 0.67 | 0.73 | 0 | Learning rate |
| | 1.0E-04 | 0.19998 | 0.729 | 0.69 | 0.78 | 1 | Learning rate |
| | 1.1E-04 | 0.19998 | 0.763 | 0.73 | 0.8 | 2 | Learning rate |
| | 1.2E-04 | 0.19998 | 0.758 | 0.71 | 0.81 | 3 | Learning rate |
| | 1.3E-04 | 0.19998 | 0.771 | 0.73 | 0.82 | 4 | Learning rate |
| | 1.4E-04 | 0.19998 | 0.761 | 0.71 | 0.82 | 5 | Learning rate |
| | 1.5E-04 | 0.19998 | 0.769 | 0.74 | 0.8 | 6 | Learning rate |
| | 1.6E-04 | 0.19998 | 0.787 | 0.74 | 0.84 | 7 | Learning rate |
| | 1.7E-04 | 0.19998 | 0.791 | 0.76 | 0.83 | 8 | Learning rate |
| | 1.8E-04 | 0.19998 | 0.792 | 0.75 | 0.84 | 9 | Learning rate |
| | 1.8E-04 | 0.19998 | 0.778 | 0.73 | 0.83 | 0 | Weight decay |
| | 1.8E-04 | 0.19999 | 0.775 | 0.73 | 0.83 | 1 | Weight decay |
| | 1.8E-04 | 0.2 | 0.79 | 0.74 | 0.85 | 2 | Weight decay |
| | 1.8E-04 | 0.20001 | 0.79 | 0.75 | 0.84 | 3 | Weight decay |
| | 1.8E-04 | 0.20002 | 0.788 | 0.75 | 0.83 | 4 | Weight decay |
| | 1.8E-04 | 0.20003 | 0.805* | 0.78 | 0.84 | 5 | Weight decay |

| | | | | | | | |
|---|---|---|---|---|---|---|---|
| | 1.8E-04 | 0.20004 | 0.778 | 0.72 | 0.84 | 6 | Weight decay |
| | 1.8E-04 | 0.19997 | 0.784 | 0.74 | 0.83 | 7 | Weight decay |
| | 1.8E-04 | 0.19996 | 0.796 | 0.77 | 0.82 | 8 | Weight decay |
| | 1.8E-04 | 0.19995 | 0.784 | 0.74 | 0.83 | 9 | Weight decay |
| | 1.8E-04 | 0.19994 | 0.787 | 0.74 | 0.84 | 10 | Weight decay |
| **Loneliness** | 1.0E-05 | 0.2 | 0.737 | 0.69 | 0.79 | 0 | Learning rate |
| | 2.0E-05 | 0.2 | 0.742 | 0.7 | 0.79 | 1 | Learning rate |
| | 3.0E-05 | 0.2 | 0.764 | 0.74 | 0.79 | 2 | Learning rate |
| | 4.0E-05 | 0.2 | 0.794 | 0.77 | 0.82 | 3 | Learning rate |
| | 5.0E-05 | 0.2 | 0.808 | 0.79 | 0.83 | 4 | Learning rate |
| | 6.0E-05 | 0.2 | 0.813 | 0.81 | 0.82 | 5 | Learning rate |
| | 7.0E-05 | 0.2 | 0.829 | 0.8 | 0.86 | 6 | Learning rate |
| | 8.0E-05 | 0.2 | 0.844 | 0.81 | 0.88 | 7 | Learning rate |
| | 9.0E-05 | 0.2 | 0.853 | 0.82 | 0.89 | 8 | Learning rate |
| | 1.0E-04 | 0.2 | 0.849 | 0.82 | 0.88 | 9 | Learning rate |
| | 9.0E-05 | 0.2 | 0.854 | 0.83 | 0.88 | 0 | Weight decay |
| | 9.0E-05 | 0.20001 | 0.866 | 0.84 | 0.89 | 1 | Weight decay |
| | 9.0E-05 | 0.20002 | 0.861 | 0.84 | 0.89 | 2 | Weight decay |
| | 9.0E-05 | 0.20003 | 0.865 | 0.84 | 0.89 | 3 | Weight decay |
| | 9.0E-05 | 0.20004 | 0.852 | 0.82 | 0.89 | 4 | Weight decay |
| | 9.0E-05 | 0.20005 | 0.87 | 0.84 | 0.9 | 5 | Weight decay |
| | 9.0E-05 | 0.20006 | 0.861 | 0.83 | 0.89 | 6 | Weight decay |
| | 9.0E-05 | 0.19999 | 0.873* | 0.84 | 0.91 | 7 | Weight decay |
| | 9.0E-05 | 0.19998 | 0.865 | 0.85 | 0.89 | 8 | Weight decay |
| | 9.0E-05 | 0.19997 | 0.861 | 0.84 | 0.89 | 9 | Weight decay |
| | 9.0E-05 | 0.19996 | 0.868 | 0.84 | 0.9 | 10 | Weight decay |
| **Mobility limitations** | 1.0E-05 | 0.2 | 0.838 | 0.86 | 0.82 | 0 | Learning rate |
| | 2.0E-05 | 0.2 | 0.85* | 0.84 | 0.86 | 1 | Learning rate |
| | 3.0E-05 | 0.2 | 0.845 | 0.83 | 0.86 | 2 | Learning rate |
| | 4.0E-05 | 0.2 | 0.832 | 0.82 | 0.85 | 3 | Learning rate |
| | 5.0E-05 | 0.2 | 0.829 | 0.82 | 0.84 | 4 | Learning rate |
| | 2.0E-05 | 0.2 | 0.831 | 0.82 | 0.84 | 0 | Weight decay |
| | 2.0E-05 | 0.20001 | 0.831 | 0.82 | 0.84 | 1 | Weight decay |
| | 2.0E-05 | 0.20002 | 0.83 | 0.82 | 0.84 | 2 | Weight decay |
| | 2.0E-05 | 0.20003 | 0.83 | 0.82 | 0.84 | 3 | Weight decay |
| | 2.0E-05 | 0.19999 | 0.825 | 0.81 | 0.84 | 4 | Weight decay |
| | 2.0E-05 | 0.19998 | 0.824 | 0.81 | 0.84 | 5 | Weight decay |
| | 2.0E-05 | 0.19997 | 0.828 | 0.81 | 0.84 | 6 | Weight decay |
| | 2.0E-05 | 0.19996 | 0.829 | 0.82 | 0.84 | 7 | Weight decay |
| | 2.0E-05 | 0.19995 | 0.828 | 0.82 | 0.84 | 8 | Weight decay |

**Supplementary Table 4.** The F1 score, precision and recall values based on optimal settings and strict evaluation matching.

| Model | Learning rate | Weight decay | F1 score | Precision | Recall |
|---|---|---|---|---|---|
| **Falling** | 0.00002 | 0.19998 | 0.775 | 0.76 | 0.79 |
| **Incontinence** | 0.00009 | 0.20001 | 0.693 | 0.65 | 0.74 |
| **Loneliness** | 0.00006 | 0.20001 | 0.801 | 0.79 | 0.81 |
| **Mobility** | 0.00002 | 0.2 | 0.842 | 0.82 | 0.86 |

**Supplementary Table 5.** Numbers of the individuals with and without the deficit and % of total in the age category identified by the named entity recognition model in the electronic health record data. We note that that total number subjects equals 99298, after excluding for subjects missing gender, birth and death year/month data.

| | Age group | N healthy | N deficit (%) |
|---|---|---|---|
| **Falls** | <60 | 28436 | 8682 (23.4) |
| | 60-70 | 25987 | 11846 (31.3) |
| | 70-80 | 11996 | 10300 (46.2) |
| | 80+ | 826 | 985 (54.3) |
| **Incontinence** | <60 | 35963 | 892 (2.4) |
| | 60-70 | 35389 | 2097 (5.6) |
| | 70-80 | 18751 | 3498 (15.7) |
| | 80+ | 1382 | 429 (23.7) |
| **Loneliness** | <60 | 36135 | 736 (2.0) |
| | 60-70 | 36438 | 1079 (2.9) |
| | 70-80 | 20690 | 1606 (7.2) |
| | 80+ | 1608 | 203 (11.2) |
| **Mobility, independent with limitations** | <60 | 33590 | 3562 (9.6) |
| | 60-70 | 31208 | 6680 (17.6) |
| | 70-80 | 14024 | 8328 (37.3) |
| | 80+ | 924 | 887 (48.8) |
| **Mobility, needs personal assistance** | <60 | 35441 | 1726 (4.6) |
| | 60-70 | 34467 | 3462 (9.1) |
| | 70-80 | 17843 | 4548 (20.3) |
| | 80+ | 1280 | 531 (29.3) |

**Supplementary Figures**

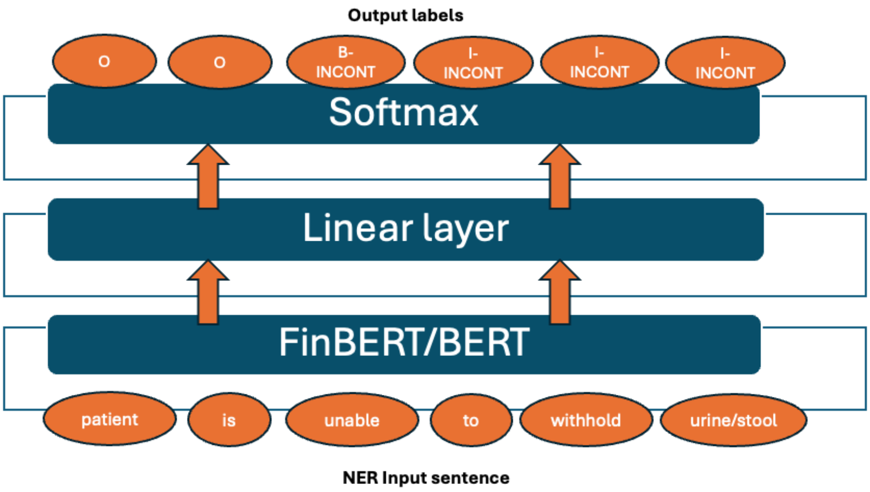


**Supplementary Figure 1.** Architecture of the named entity recognition task using the FinBERT/BERT model and BIO tagging (Incontinence shorten to INCONT) with an example passage of incontinence (translated from Finnish for readability). NER, named entity recognition.

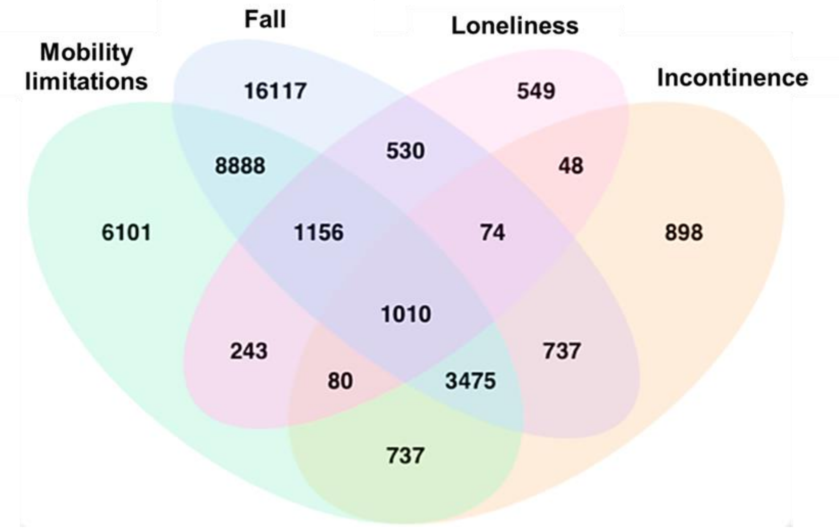


**Supplementary Figure 2**. The overlap of the conditions among the patients identified by the named entity recognition model.
